# Real-world systemic therapy utilization and survival in synchronous metastatic solid cancer: a comprehensive nationwide analysis

**DOI:** 10.64898/2026.07.20.26358468

**Authors:** Ellis Slotman, Lieke M. van Disseldorp, Gwen de Jong, Heidi Fransen, Anna K. L. Reyners, Jolien Tol, Agnes Jager, Hans M. Westgeest, Gabe S. Sonke, Hanneke W. M. van Laarhoven, Lia van Zuylen, Michel van den Heuvel, Miriam Koopman, Egbert Smit, Natasja Raijmakers, Sabine Siesling

## Abstract

**Introduction:** This study aimed to provide population-level survival trends during the era in which new systemic therapies transformed treatment guidelines for metastatic cancer, as well as insights on the real-world use of these treatments and associated survival.

**Methods:** Adults diagnosed with synchronous metastatic solid cancer in 2008–2022 (22 cancer types) were identified from the Netherlands Cancer Registry. Median overall survival (OS) was assessed by five-year diagnostic period. For 2018-2022, systemic therapy use in any treatment line was analyzed and survival percentiles within treatment and cancer types were estimated with Kaplan-Meier survival analyses.

**Results:** Median OS in the overall cohort (n=280,419 patients) improved from 6 to 8 months between 2008–2012 and 2018–2022. Among patients diagnosed in 2018–2022, 15% received immunotherapy, 15% targeted therapy, 29% chemotherapy and/or traditional hormone therapy only, and 39% no systemic therapy. In some cancer types, a relatively large proportion of treated patients had long-term survival (e.g., immunotherapy in melanoma: p50 = 67 months). Other cancer types had a smaller subset of treated patients (p10 and p25) with substantially better outcomes than the median (e.g., targeted therapy in NSCLC: p50 = 22 months, p10 = 96 months).

**Conclusion:** Population-level survival for patients with synchronous metastatic solid cancer has modestly improved over time. The marked survival heterogeneity within cancer and treatment types highlights both the potential and uncertainty associated with (novel) treatments. Improved prediction of treatment effects and clear communication regarding survival expectation remain critical. Presenting multiple survival scenarios over median survival alone can support decision making.

## Introduction

Globally, approximately 20 million people are diagnosed with cancer each year.^1^ In around one fifth of patients in the Netherlands, the cancer has already metastasized at diagnosis (synchronous metastatic cancer), although this proportion differs considerably between cancer types.^2^ Metastatic cancer is typically incurable.

Recently, the treatment landscape for metastatic cancer has changed drastically. Novel types of systemic treatment, including immunotherapies and targeted therapies, can significantly improve life expectancy.^3–8^ They differ from traditional systemic treatments (chemotherapy and traditional hormonal therapy), as they employ novel targeted mechanisms such as immune checkpoint inhibition, blockade of specific molecular pathways, next-generation hormone receptor targeting or targeted radiopharmaceutical therapy.^9–12^ Additionally, they can induce different typical illness trajectories, including short-lived improvement with targeted therapy and long-term ongoing responses in immunotherapy.^8^

A nationwide Dutch study analyzing population-level survival trends for all patients with synchronous metastatic cancer through 2018 found only marginal survival improvements for most cancer types despite the introduction of new medicines.^13^ However, in clinical practice, only a subset of patients is eligible for these novel systemic treatments.^14,15^ As a result, population-level survival trends including those who did not receive (novel) treatments mask meaningful survival improvements in specific subgroups. Therefore, more detailed analyses on survival outcomes within treatment subgroups enhance insight in real-world outcomes of anti-cancer treatments for metastatic cancer. Moving beyond median survival by evaluating a range of survival scenarios (percentiles) shows the variability of treatment outcomes in routine practice, which can inform more realistic treatment expectations. Additionally, treatment options have further expanded since 2018, with many new drugs and indications predominantly for metastatic cancer.^16,17^ Moreover, longer follow-up can now capture effects of earlier innovations. Examining more recent survival trends is therefore warranted.

The aim of our study was to compare overall survival (OS) among patients diagnosed with synchronous metastatic cancer in 2018-2022 with survival outcomes from earlier periods (2008-2012 and 2013-2017) in the Netherlands. Moreover, we aimed to describe real-world utilization of novel and traditional systemic therapies covering multiple treatment lines, and to analyze a range of survival scenarios among systemically treated patients for more granular insight into survival outcomes in daily practice.

## Methods

### Study population and data sources

For this observational cohort study, all adult patients diagnosed with synchronous metastatic solid cancer in the period 2008-2022 were selected from the nationwide Netherlands Cancer Registry (NCR). The NCR includes all newly diagnosed malignancies since 1989, mainly based on notification by the National Pathology Archive (Palga).^18^ The NCR registers stage at diagnosis, but metachronous metastases are not routinely registered so this study only includes patients with synchronous metastatic disease. Patient and disease characteristics, as well as data on initial treatment, were retrieved from the NCR. Data on vital status were obtained through annual linkage with the National Municipal Personal Records Database and were available until January 31, 2026.

In the NCR, treatment data are generally limited to the initial (first-line) treatment. To facilitate analyses of utilization and survival outcomes of systemically treated patients diagnosed in the most recent cohort (2018–2022), we supplemented the NCR treatment data with information from the Dutch National Hospital Care Registration (LBZ).^19^ The LBZ, hosted by Dutch Hospital Data (DHD), receives medical data from all Dutch hospitals. The LBZ data in our study consisted of data on systemic therapies administered in 2018-2023, covering initial and subsequent systemic treatments. In the LBZ systemic treatment use was identified from treatment procedure data as well as data on add-on medications (high-cost drugs that are reimbursed separately from other hospital care in the Netherlands). For this study, patients who were diagnosed or treated in hospitals that did not consent to data sharing (20%), whose records in the NCR could not be successfully linked to the LBZ (1%), or who were diagnosed with metastatic cancer at death were excluded.

This study was approved by the Scientific Committee and Privacy Review Board of the NCR (reference number K23.201).

### Definitions

Synchronous metastatic cancer was defined as any distant metastasis at diagnosis, as defined by 1) M1 disease according to the TNM classification, or 2) distant metastases according to the Extent of Disease classification system, or 3) a tumor of unknown primary (C80.9).

Patients were categorized into 22 cancer types based on primary cancer site and morphology (Supplementary table 1). Cancers with <200 metastatic cancer diagnoses in a 5-year diagnostic period, as well as poorly defined categories (e.g., not-otherwise specified lung cancer) were excluded. Moreover, patients with multiple primary metastatic tumors diagnosed within the study period were excluded.

For the analyses of patients diagnosed in 2018-2022 who received systemic treatment, patients were categorized into four treatment groups based on all (both initial and subsequent) systemic treatments received. Groups 1-3 included patients who received novel types of systemic treatment: 1) immunotherapy, 2) novel targeted therapy, 3) radionuclides and/or, specifically for prostate cancer, androgen receptor–targeted agents (ARTAs, hereafter referred to as novel hormonal agents). Included therapies in each treatment group are presented in Supplementary Table 2. These groups are non-mutually exclusive, meaning that patients could be included in more than one category if they received multiple novel treatments (e.g., patients treated with both immunotherapy and targeted therapy within the same or subsequent treatment lines were included in both category 1 and 2). Patients in groups 1–3 could also have received traditional systemic therapies (chemotherapy or traditional hormone therapy) in addition to the novel type of systemic treatment. Group 4 included patients who received traditional systemic therapy only and did not receive any of the novel systemic treatments from categories 1–3. Local immunotherapy or local chemotherapy were not included in the analyses. Categorization of patients into the treatment groups was based the combination of NCR and LBZ data.

### Statistical analyses

Patients’ characteristics at diagnosis were descriptively analyzed by cancer type and period of diagnosis (2008-2012, 2013-2017, and 2018-2022). Median OS was evaluated for each of the three time periods for all patients diagnosed with synchronous metastatic cancer (regardless of whether they received treatment or the type of treatment), both for the total cohort and stratified by cancer type. OS was defined as the number of months between date of diagnosis and death or end of follow-up.

For patients diagnosed in 2018-2022 who had received systemic treatment, more detailed analyses were conducted. Utilization of systemic treatment was analyzed as the proportion of patients in each treatment group, for the total cohort and per cancer type. Furthermore, for each treatment group, the Kaplan–Meier method was used to determine OS for the total cohort and per cancer type, calculating the following survival percentiles (representing scenarios): 10th (best-case), 25th (upper-typical), 50th (median), 75th (lower-typical), and 90th (worst-case). These percentiles represent the number of months survived by 10%, 25%, 50%, 75% and 90% of patients, respectively (Figure 1). The survival scenarios and terminology are consistent with previous research.^20–22^ Percentiles were not reported if there was insufficient follow-up time to calculate point estimates. Median follow-up time was calculated using the reverse Kaplan-Meier method.

**Figure 1:**
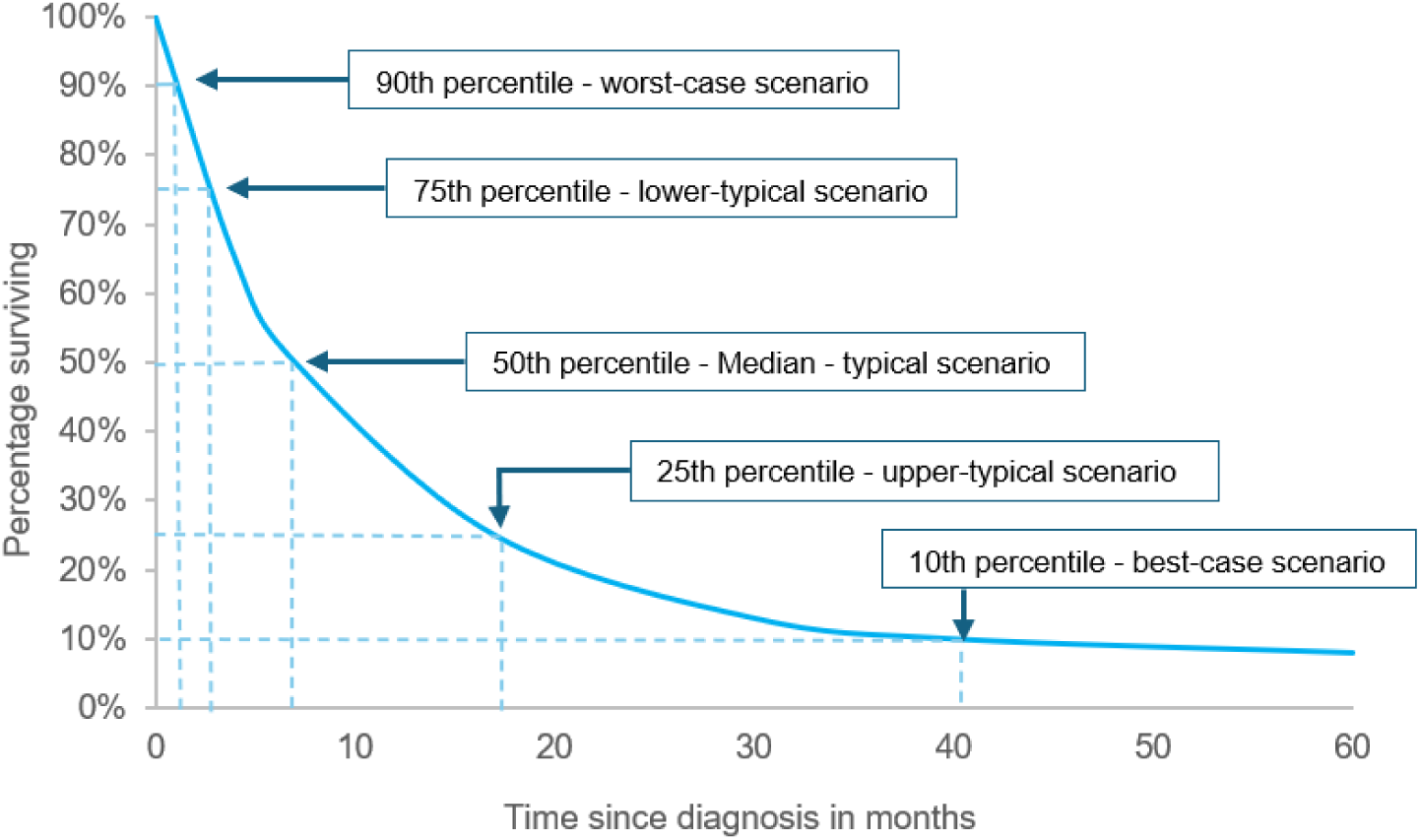
Illustration of the survival curve percentiles and the corresponding survival scenarios.

Two sensitivity analyses were conducted to minimize the impact of immortal time bias and the impact of combination therapies on the survival scenarios. The first sensitivity analysis included only patients who received one or more types of novel systemic anti-cancer therapies (1) immunotherapy, 2) targeted therapy, and/or 3) radionuclides and/or novel hormonal agents) as *initial treatment*, thereby reducing immortal time bias from therapies administered in later lines of therapy. The second sensitivity analysis included only patients who received one or more medicines from a *single* type of novel systemic anticancer therapy (1) immunotherapy, 2) targeted therapy, or 3) radionuclides, or 4) novel hormonal agents), excluding those treated with more than one of these categories. This was done to obtain insight into the survival outcomes for each specific type of novel systemic therapy separately. In both sensitivity analyses, patients receiving novel systemic therapies may, in addition, also have received traditional systemic therapy (chemotherapy or traditional hormone therapy), as this reflects real-world practice in which novel and traditional treatments are often combined or given sequentially.

In all survival analyses, treatment groups with fewer than 50 patients were excluded from presentation. Analyses were performed using Stata software version 17.

## Results

In total, 280,419 patients diagnosed with synchronous metastatic cancer between 2008 and 2022 were included in this study (2008-2012: n=85,295, 2013-2017: n=92,177, 2018-2022: n=102,947) (Table 1). Median age at diagnosis in the total cohort increased from 69 years (2008-2012) to 71 years (2018-2022), with the highest increase for melanoma. The percentage of female patients was stable in the total cohort but changed to some extent across cancer types.

**Table 1:** Number, median age and % female of all patients diagnosed with synchronous metastatic cancer between 2008 and 2022 per cancer type and period of diagnosis (n=85,295).

|  | 2008-2012 |  |  | 2013-2017 |  |  | 2018-2022 |  |  |
| --- | --- | --- | --- | --- | --- | --- | --- | --- | --- |
|  | N | Median age (IQR) | % female | N | Median age (IQR) | % female | N | Median age (IQR) | % female |
| Total cohort | 85295 | 69 (61-77) | 42 | 92177 | 69 (61-77) | 43 | 102947 | 71 (63-78) | 42 |
| Bile duct cancer | 825 | 70 (61-78) | 49 | 1364 | 68 (60-76) | 49 | 1906 | 70 (63-77) | 50 |
| Bladder cancer | 1625 | 73 (63-80) | 31 | 2047 | 72 (64-79) | 32 | 2841 | 73 (67-80) | 33 |
| Breast cancer | 3389 | 64 (51-77) | 99 | 4196 | 64 (52-76) | 99 | 4882 | 65 (52-75) | 99 |
| Cervical cancer | 349 | 59 (49-74) | 100 | 394 | 59 (49-73) | 100 | 338 | 56 (48-70) | 100 |
| Colon cancer | 10631 | 70 (62-78) | 47 | 11112 | 70 (61-78) | 46 | 10331 | 72 (61-79) | 49 |
| Endometrial cancer | 561 | 69 (63-78) | 100 | 814 | 70 (65-77) | 100 | 1033 | 72 (64-77) | 100 |
| Esophageal/cardia cancer | 4688 | 67 (59-75) | 22 | 5031 | 68 (61-75) | 23 | 5459 | 70 (62-76) | 23 |
| Gallbladder cancer | 373 | 72 (63-79) | 72 | 412 | 71 (64-78) | 73 | 570 | 72 (64-79) | 66 |
| Gastric cancer | 2714 | 70 (62-78) | 40 | 2540 | 72 (63-78) | 39 | 3236 | 71 (61-78) | 39 |
| HNSCC | 262 | 64 (59-71) | 27 | 346 | 67 (61-73) | 27 | 416 | 69 (61-75) | 25 |
| Hepatocellular cancer | 380 | 67 (59-75) | 21 | 553 | 69 (62-76) | 23 | 721 | 72 (65-77) | 23 |
| Kidney cancer | 2506 | 68 (60-76) | 35 | 2528 | 69 (60-76) | 34 | 2607 | 70 (62-77) | 32 |
| Melanoma | 1006 | 62 (51-72) | 41 | 1138 | 65 (54-73) | 39 | 1525 | 68 (57-76) | 36 |
| NET | 1022 | 67 (59-75) | 48 | 1262 | 67 (59-74) | 48 | 1628 | 68 (59-75) | 44 |
| NSCLC | 20933 | 67 (59-75) | 40 | 23428 | 68 (60-75) | 43 | 25512 | 69 (62-75) | 45 |
| Ovarian cancer | 1465 | 69 (61-77) | 100 | 1715 | 70 (61-77) | 100 | 2209 | 71 (62-78) | 100 |
| Pancreatic cancer | 5868 | 69 (62-77) | 48 | 7144 | 70 (63-77) | 47 | 8434 | 72 (64-78) | 49 |
| Prostate cancer | 6848 | 74 (67-80) | 0 | 8462 | 74 (67-80) | 0 | 13684 | 74 (68-80) | NA |
| Rectal cancer | 4110 | 67 (59-75) | 38 | 4224 | 67 (59-75) | 38 | 3402 | 68 (57-77) | 38 |
| SCLC | 5823 | 67 (61-74) | 42 | 5887 | 68 (62-74) | 47 | 5683 | 69 (63-75) | 50 |
| Sarcoma | 644 | 62 (49-74) | 45 | 664 | 66 (53-76) | 42 | 750 | 66 (52-75) | 40 |
| Cancer of unknown primary | 9273 | 75 (65-82) | 51 | 6916 | 74 (66-82) | 49 | 5780 | 76 (68-83) | 49 |

### Median overall survival in all patients with synchronous metastatic cancer

Median OS for the total cohort increased from 6 months in 2008-2012 to 8 months in 2018-2022 (Figure 2). For most cancer types, median OS remained unchanged across the three time periods. Patients with hepatocellular, pancreatic cancer, and cancer of unknown primary had the poorest survival overall, with median OS of 2 months across all periods and 1 month for cancer of unknown primary in 2018–2022. The most pronounced improvements in median OS were observed in metastatic breast cancer (11 months), melanoma (17 months), and prostate cancer (17 months).

**Figure 2:**
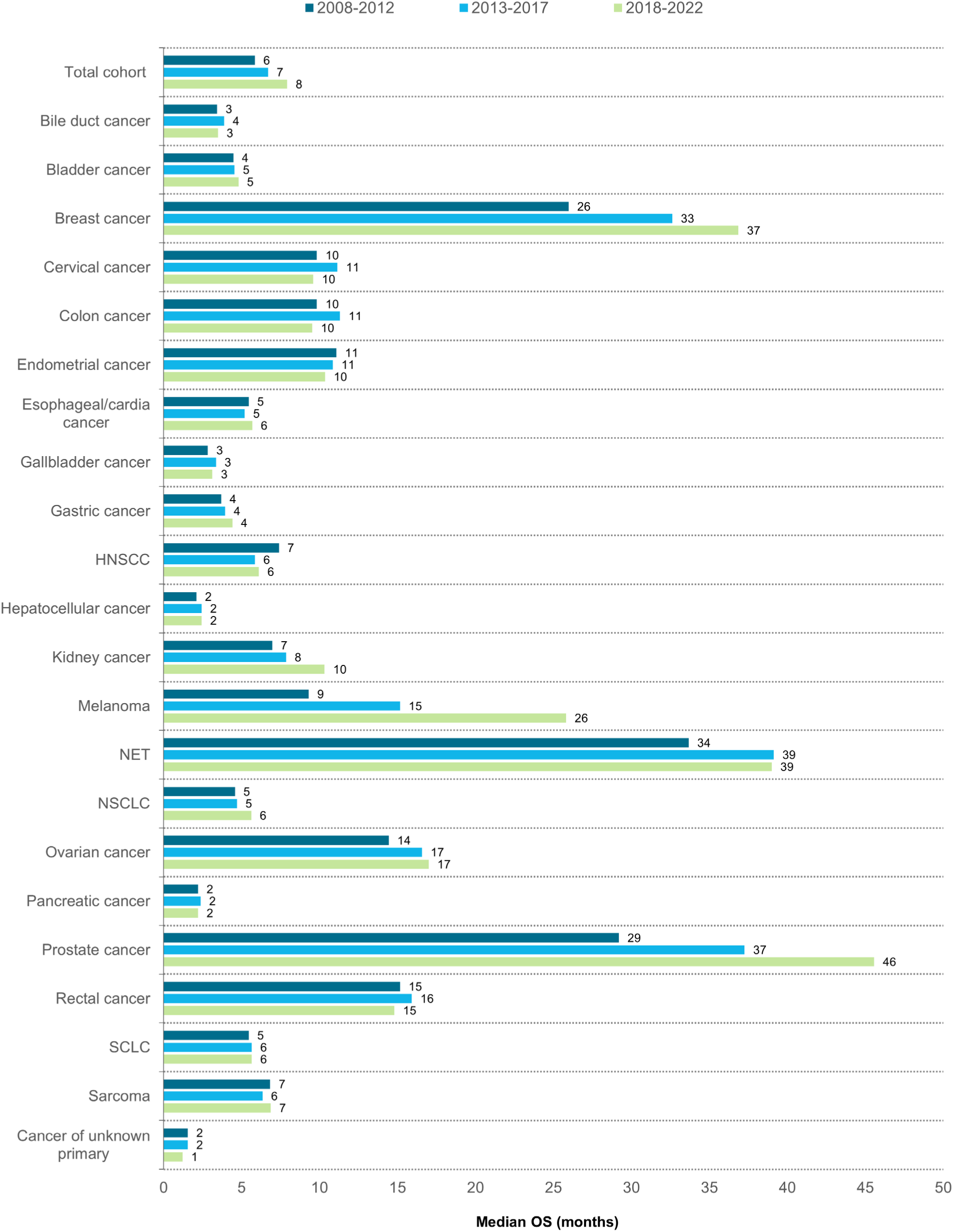
Median overall survival of patients diagnosed with synchronous metastatic cancer per cancer type and period of diagnosis (2008-2012, 2013-2017, 2018-2022).

### Utilization of systemic treatment

Of all patients diagnosed with synchronous metastatic cancer in the most recent period (2018-2022), 82,544 (80%) could be linked to the LBZ to retrieve information on systemic therapy in all treatment lines. The median age and sex distributions of these patients were comparable to those of the total cohort diagnosed in 2018-2022 (Supplementary Table 3).

Among these patients, 15% received immunotherapy, with the highest use in melanoma (64%), kidney cancer (44%), and NSCLC (43%) (Table 2). Targeted therapy was administered to 15% of all patients, with the highest use in breast cancer (45%), rectal cancer (43%), and kidney cancer (38%). Therapies classified as radionuclides or novel hormonal agents were used in prostate cancer (36%) and NET (13%). Traditional systemic therapy only was administered to 29% of all patients, most frequently in small-cell lung cancer (68%), endometrial cancer (64%), and prostate cancer (59%). In total, 39% of all patients did not receive any of the aforementioned systemic therapies.

**Table 2:** Utilization of systemic therapies in patients diagnosed with synchronous metastatic cancer between 2018-2022 (n=82544).

|  | Immunotherapy* | Targeted therapy* | Radionuclides and/or novel hormonal agents* | Traditional systemic therapy only (chemotherapy and/or traditional hormone therapy) |
| --- | --- | --- | --- | --- |
|  | N (%) | N (%) | N (%) | N (%) |
| Total cohort (N=82544) | 12069 (15) | 12117 (15) | 4125 (5) | 23903 (29) |
| Bile duct cancer (N=1556) | x | 10 (1) | x | 556 (36) |
| Bladder cancer (N=2235) | 405 (18) | 31 (1) | x | 571 (26) |
| Breast cancer (N=3942) | 74 (2) | 1757 (45) | x | 1750 (44) |
| Cervical cancer (N=298) | 19 (6) | 83 (28) | x | 82 (28) |
| Colon cancer (N=8264) | 147 (2) | 3064 (37) | x | 1567 (19) |
| Endometrial cancer (N=857) | 21 (2) | 23 (3) | x | 552 (64) |
| Esophageal/cardia cancer (N=4513) | 143 (3) | 684 (15) | x | 1580 (35) |
| Gallbladder cancer (N=474) | x | x | x | 176 (37) |
| Gastric cancer (N=2635) | 54 (2) | 369 (14) | x | 844 (32) |
| HNSCC (N=383) | 69 (18) | 34 (9) | x | 32 (8) |
| Hepatocellular cancer (N=597) | 59 (10) | 160 (27) | x | x |
| Kidney cancer (N=2231) | 991 (44) | 846 (38) | x | 12 (1) |
| Melanoma (N=1369) | 880 (64) | 389 (28) | x | x |
| NET (N=1405) | 11 (1) | 89 (6) | 189 (13) | 716 (51) |
| NSCLC (N=20772) | 8855 (43) | 2524 (12) | x | 1961 (9) |
| Ovarian cancer (N=1600) | 37 (2) | 474 (30) | x | 764 (48) |
| Pancreatic cancer (N=6499) | 29 (0) | 29 (0) | x | 2024 (31) |
| Prostate cancer (N=10726) | 47 (0) | 171 (2) | 3899 (36) | 6316 (59) |
| Rectal cancer (N=2824) | 15 (1) | 1214 (43) | x | 686 (24) |
| SCLC (N=4566) | 116 (3) | 30 (0) | x | 3105 (68) |
| Sarcoma (N=542) | x | 69 (13) | x | 156 (29) |
| Cancer of unknown primary (N=4256) | 81 (2) | 66 (2) | x | 439 (10) |

\* - Categories are not mutually exclusive. Only the category 'traditional systemic therapy' contains only patients who did not receive any treatments from the other groups.
x – Data not shown because this treatment type was not administered for this cancer type, or because the number of treated patients was <10.
*Note: This table only includes patients who have received systemic therapy. Patients who received only local treatment or no anti-cancer treatment are not included.*

### Survival scenarios in patients receiving systemic treatment

For the total cohort of patients receiving *immunotherapy* (n=12,069), median OS was 17 months, ranging from 3 months (p90) to 46 months (p25) (Figure 3A). Survival differences between p90, p75, and the median were generally small, except for melanoma (median OS 67 months vs. p75 14 months and p90 6 months). The largest differences between the median and the p25 were observed for breast cancer, kidney cancer and NSCLC, ranging from 24 months in NSCLC to 30 months in kidney cancer. The largest difference between the median and the best-case scenario (p10) was 34 months for hepatocellular cancer. In most cancer types, the p10 was not reached.

**Figure 3:**
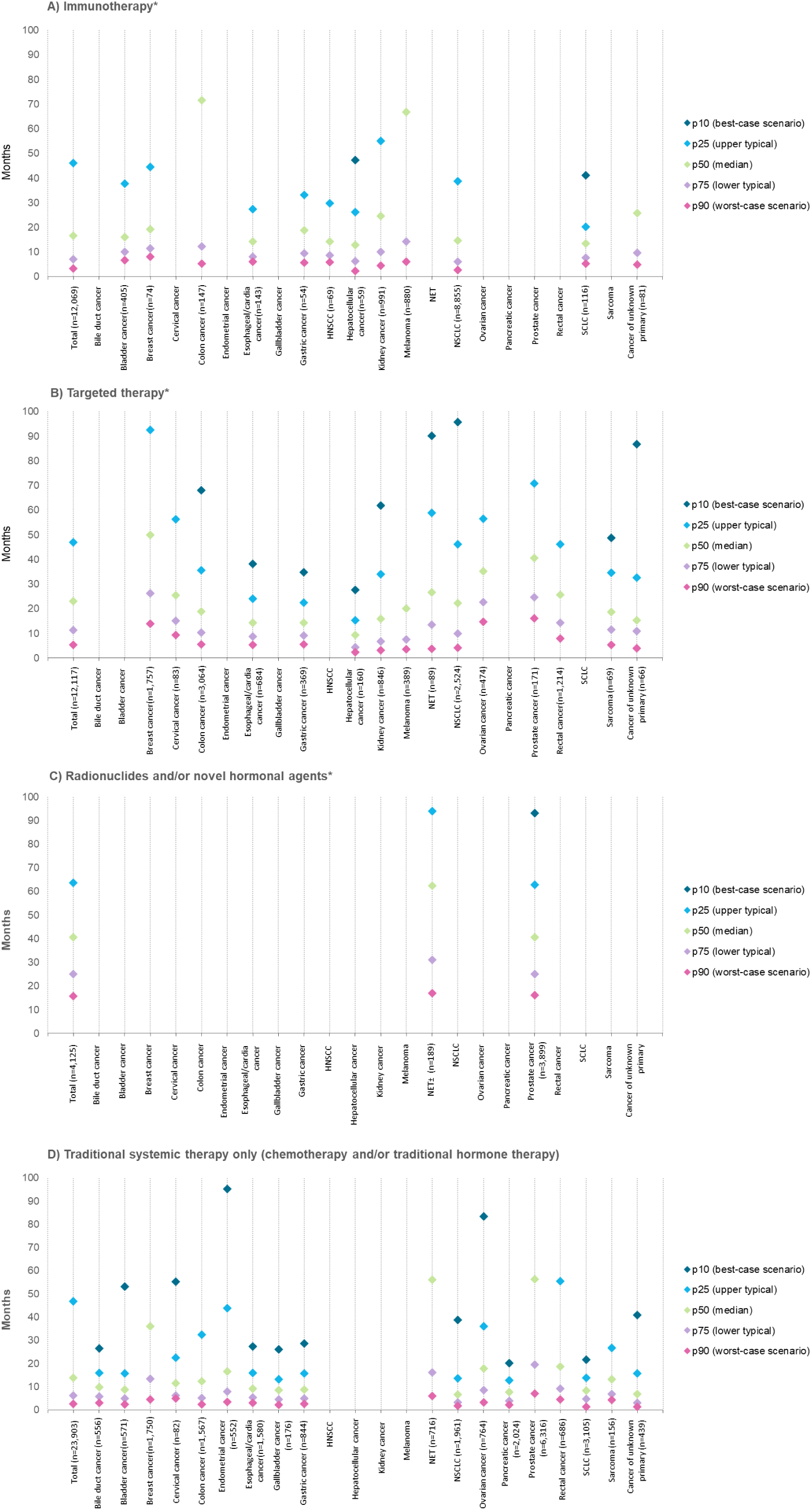
Survival scenarios in patients diagnosed with synchronous metastatic cancer in **2018-2022** and treated with novel systemic anticancer therapies or traditional systemic therapy only. *Categories are not mutually exclusive. ± In NET only radionuclides were administered. Notes: - Systemic treatment data were available from January 1, 2018, until December 31, 2023. - For some treatment groups and cancer types, the highest percentiles cannot be reported due to insufficient follow-up time to calculate point estimates. - Treatment groups with fewer than 50 patients were excluded from presentation.

For the total cohort of patients receiving *targeted therapy* (n=12,117), median OS was 23 months, ranging from 5 months (p90) to 47 months (p25) (Figure 3B). The largest differences between the median and the highest reached survival percentile were observed in breast cancer (p25; 43-month difference) and in NSCLC (p10; 73-month difference). The smallest differences between the median and the best-case scenario (p10) were observed in patients with hepatocellular cancer (18 months), gastric cancer (20 months), and esophagus/cardia cancer (24 months).

For *radionuclides and/or novel hormonal agents,* the total cohort (n=4,125) had a median survival of 41 months, ranging from 16 months (p90) to 64 months (p25) (Figure 3C). For NET, the difference between the median and the upper-typical scenario (p25) was 32 months, and the best-case scenario was not reached; for prostate cancer, the difference between the median and the best-case scenario (p10) was 53 months.

For patients receiving *traditional systemic therapy only* (n=23,903), survival times were 14 months (median), 3 months (p90), and 47 months (p25), respectively (Figure 3D). The dispersion between the survival scenarios was generally smaller compared to the novel systemic therapies.

Sensitivity analyses showed similar patterns in survival outcomes, with consistent dispersion across survival percentiles (Supplementary Figures 1 and 2). Median follow-up times for the total cohort as well as per cancer type are reported in Supplementary Table 4.

## Discussion

In this nationwide study, we evaluated real-world trends in survival among patients diagnosed with synchronous metastatic cancer in 2008-2022 and examined utilization and survival outcomes of novel and traditional types of systemic therapies for patients diagnosed in 2018– 2022.

Median OS for the total population improved from six months (2008-2012) to eight months (2018-2022). One likely explanation for the relatively limited improvement is the utilization of novel systemic therapies; 15% of all patients received immunotherapy and 15% targeted therapy, while many patients received traditional systemic treatments only (29%) or no systemic treatment (39%). Although therapeutic options have expanded considerably in recent years, their availability remains confined to specific cancer types and patient subgroups. In addition, in routine practice factors such as poor performance status or comorbidities may limit eligibility or be a reason to forego treatment in shared decision-making. Consequently, potential benefits experienced by small subgroups receiving systemic therapies do not translate into improvements in the population-level median survival trend. In addition, Improved staging, including advanced imaging, may also influence survival trends by detecting previously unrecognized metastases.^23–25^ In addition, increasing age at diagnosis across several cancer types, likely reflecting population ageing^26^, may affect OS and treatment eligibility through higher comorbidity and frailty. ^27,28^

Despite the limited survival improvement in the total cohort, this study demonstrates meaningful progress in specific patient populations. In metastatic breast cancer, melanoma and prostate cancer, population-level median OS increased substantially over time. Simultaneously, a high utilization of (novel) systemic treatments was observed in these cancer types and a relatively large proportion of treated patients achieved relatively long-term OS. For example, half of patients survived at least 50 months with targeted therapy for breast cancer and at least 67 months with immunotherapy for melanoma. In other cancer types, such as NSCLC, kidney cancer and colon and rectal cancer, population-level median OS remained largely unchanged, despite relatively high systemic therapy use. Although we did observe long-term survival outcomes with novel systemic treatments within these cancer types, these were generally more confined to the upper-typical and best-case scenarios. The variability in the proportion of patients experiencing long-term survival is likely strongly influenced by differences in treatment response rates across cancer types, as previously shown for anti-PD-1/PD-L1 immunotherapy^29^ and BRAF-inhibitor therapies^30^, as well as by heterogeneity in therapeutic resistance mechanisms within and between cancer types.^31^ Moreover, (promising) novel therapies were introduced in the Netherlands during and after the study period. This highlights the need for continuous monitoring of utilization and survival trends.

Overall, the findings highlight both the promise and the uncertainty of novel systemic treatments. While some treated patients achieve long-term survival, others have only short survival times despite being treated with these therapies. This underscores the importance of communicating this uncertainty in clinical practice, as it has been associated with increased anxiety, distress, and disappointment when treatment response is insufficient.^32,33^ Mixed framing, in which best-case, worst-case and typical outcomes are discussed in clinical conversations can support nuanced discussions of treatment options.^34,35^ It has also been shown that patients prefer multiple survival scenarios over presentation of median survival alone.^36,37^ Moreover, timely integration of palliative care alongside systemic treatment is important, as it can support prognostic understanding and navigate prognostic uncertainty, help manage symptoms, toxicities and psychological distress, and assist in advance care planning discussions.^38,39^

Predicting response and resistance to systemic therapies remains challenging.^40–43^ Since efficacy is often established in selected trial populations, use in broader real-world populations may lead to more variable and potentially lower response and survival.^44–46^ Furthermore, often considerable uncertainty exists at the time of regulatory approval as well, since an increasing number of therapies are introduced based on small, often single-arm studies and surrogate endpoints, while overall survival benefits are more uncertain. Improving prediction of treatment response (e.g., through patient characteristics and/or biomarkers) and assessing real-world treatment use and outcomes in broader populations, including post-approval studies, are essential to support individualized decision making for systemic therapies. Beyond survival, pre- and post-registration studies should also include patient-reported outcomes, treatment-related toxicities and quality-of-life measures, as these are crucial for understanding the impact of these therapies.

A major strength of this study is its nationwide scope and linked datasets, which enabled assessment of systemic therapy use across multiple treatment lines. However, several limitations should be acknowledged. First, data on systemic treatments use of the patients diagnosed in 2018-2022 were available through 2023. Consequently, patients initiating a certain treatment after 2023 could not be classified accordingly, potentially leading to underestimation of the proportion of patients in each category. Furthermore, as vital status data were available through January 2026, the shorter follow-up for patients diagnosed toward the end of the study period hindered the estimation of the highest survival percentiles. In addition, some cancer types had relatively small sample sizes, which may reduce the precision of particularly the p10 and p90 percentiles. Furthermore, patients with metachronous metastases were not included. The generalizability of the findings to this group may be limited, as prior curative-setting treatments may affect eligibility for and effectiveness of systemic therapies in the metastatic setting.^47^ Furthermore, linkage to the LBZ was not possible for patients who received care in hospitals that did not consent to share data, and treatment data may be incomplete for patients who were later referred to these hospitals. Nevertheless, LBZ data covered 80% of patients diagnosed in 2018–2022; linked patients were comparable to the overall population, and the included hospitals represented university medical centers, top clinical hospitals and general hospitals, and were well-distributed across the Netherlands.

### Conclusion

Although median OS for patients with synchronous metastatic cancer showed modest improvement between 2008 and 2022, some cancer types, including melanoma, prostate, and breast cancer, showed substantial gains. The use of novel systemic therapies among patients diagnosed in 2018-2022 remained limited in the total population. Among those who received (novel) systemic therapies, we did observe long-term survival for subsets of patients, but survival outcomes were highly variable. Improving the prediction of individual outcomes and enhancing prognostic communication, including discussion of different survival scenarios, should therefore remain priorities for future research and clinical practice.

## Data Availability

The data that support the findings of this study are available from the Netherlands Comprehensive Cancer Organisation (IKNL) and Dutch Hospital Data (DHD). Data are used under licence for this study and therefore not publicly available. (Analytical) code(s) are available upon (reasonable) request in consultation with the corresponding author.

## References

1. Bray F, Laversanne M, Sung H, et al. Global cancer statistics 2022: GLOBOCAN estimates of incidence and mortality worldwide for 36 cancers in 185 countries. CA Cancer J Clin. 2024;74(3):229–263.

2. Integraal Kankercentrum Nederland (IKNL). Uitgezaaide kanker 2025: cijfers, inzichten en uitdagingen. 2025. https://iknl.nl/getmedia/0bb2f9fc-aa08-4693-8b54-0bb750e51bef/rapport-uitgezaaide-kanker-2025-definitief2.pdf

3. Palumbo MO, Kavan P, Miller WH, Jr., et al. Systemic cancer therapy: achievements and challenges that lie ahead. Front Pharmacol. 2013;4:57. doi:10.3389/fphar.2013.00057

4. Arbour KC, Riely GJ. Systemic therapy for locally advanced and metastatic non–small cell lung cancer: a review. JAMA. 2019;322(8):764–774.

5. Powers E, Karachaliou GS, Kao C, et al. Novel therapies are changing treatment paradigms in metastatic prostate cancer. J Hematol Oncol. 2020;13:1–13.

6. Ugurel S, Röhmel J, Ascierto PA, et al. Survival of patients with advanced metastatic melanoma: the impact of novel therapies. Eur J Cancer. 2016;53:125–134.

7. Jawed I, Wilkerson J, Prasad V, Duffy AG, Fojo T. Colorectal cancer survival gains and novel treatment regimens: a systematic review and analysis. JAMA oncology. 2015;1(6):787–795.

8. Geijteman ECT, Kuip EJM, Oskam J, Lees D, Bruera E. Illness trajectories of incurable solid cancers. BMJ. 2024;384:e076625. doi:10.1136/bmj-2023-076625

9. Rezazadeh-Gavgani E, Majidazar R, Lotfinejad P, Kazemi T, Shamekh A. Immune Checkpoint Molecules: A Review on Pathways and Immunotherapy Implications. Immun Inflamm Dis. Apr 2025;13(4):e70196. doi:10.1002/iid3.70196

10. Gill MR, Falzone N, Du Y, Vallis KA. Targeted radionuclide therapy in combined-modality regimens. The Lancet Oncology. 2017;18(7):e414–e423. doi:10.1016/S1470-2045(17)30379-0

11. Abah MA, Oladosu MA, Nnaemeka NJ, Agida OD. Targeted Therapeutics for Cancer Treatment: A Review of Kinase Inhibitors, Angiogenesis Inhibitors, and Other Molecularly targeted Agents.

12. Rice MA, Malhotra SV, Stoyanova T. Second-generation antiandrogens: from discovery to standard of care in castration resistant prostate cancer. Front Oncol. 2019;9:801.

13. Luyendijk M, Visser O, Blommestein HM, et al. Changes in survival in de novo metastatic cancer in an era of new medicines. JNCI: Journal of the National Cancer Institute. 2023;doi:10.1093/jnci/djad020

14. Haslam A, Prasad V. Estimation of the Percentage of US Patients With Cancer Who Are Eligible for and Respond to Checkpoint Inhibitor Immunotherapy Drugs. JAMA Netw Open. May 3 2019;2(5):e192535. doi:10.1001/jamanetworkopen.2019.2535

15. Marquart J, Chen EY, Prasad V. Estimation of the Percentage of US Patients With Cancer Who Benefit From Genome-Driven Oncology. JAMA Oncol. Aug 1 2018;4(8):1093–1098. doi:10.1001/jamaoncol.2018.1660

16. Grössmann N, Wolf S, Rothschedl E, Wild C. Twelve years of European cancer drug approval-a systematic investigation of the ‘magnitude of clinical benefit’. ESMO Open. Jun 2021;6(3):100166. doi:10.1016/j.esmoop.2021.100166

17. Cramer A, Sørup FKH, Christensen HR, Petersen TS, Karstoft K. Cancer drug applications to the EMA and the FDA: A comparison of new drugs and extension of indication in terms of approval decisions and time in review. Br J Clin Pharmacol. May 2025;91(5):1431–1438. doi:10.1111/bcp.16391

18. Integraal Kankercentrum Nederland (IKNL). Nederlandse Kankerregistratie. Accessed January 15, 2026. https://iknl.nl/nkr

19. Dutch Hospital Data. Registratie en data. https://www.dhd.nl/producten-diensten/registratie-data

20. Pape MMM. Beyond Median Overall Survival: Estimating Trends for Multiple Survival Scenarios in Patients With Metastatic Esophagogastric Cancer. JNCCN Journal of the National Comprehensive Cancer Network. 2022;20(12):1321–1329.

21. Hamers PAH, Elferink MAG, Stellato RK, et al. Informing metastatic colorectal cancer patients by quantifying multiple scenarios for survival time based on real-life data. Int J Cancer. Jan 15 2021;148(2):296–306. doi:10.1002/ijc.33200

22. Kuijper SC, Gehrels AM, van der Geest LG, et al. Survival scenarios of patients with localized and metastatic pancreatic adenocarcinoma: A population-based study. Int J Cancer. 2025/05/01 2025;156(9):1726-1735. 10.1002/ijc.35267

23. Yu B, Zhu X, Liang Z, Sun Y, Zhao W, Chen K. Clinical usefulness of (18)F-FDG PET/CT for the detection of distant metastases in patients with non-small cell lung cancer at initial staging: a meta-analysis. Cancer Manag Res. 2018;10:1859–1864. doi:10.2147/cmar.S155542

24. Vogsen M, Jensen JD, Christensen IY, et al. FDG-PET/CT in high-risk primary breast cancer-a prospective study of stage migration and clinical impact. Breast Cancer Res Treat. Jan 2021;185(1):145–153. doi:10.1007/s10549-020-05929-3

25. Heesterman BL, Peters M, Oprea-Lager DE, et al. Increased incidence of primary metastatic prostate cancer in the era of PSMA PET/CT: a population-based analysis. Eur J Nucl Med Mol Imaging. Dec 2025;53(1):350–361. doi:10.1007/s00259-025-07431-8

26. Statistics Netherlands (CBS). Elderly people. Accessed January 16, 2026. https://www.cbs.nl/en-gb/visualisations/dashboard-population/age/elderly-people

27. George M, Smith A, Sabesan S, Ranmuthugala G. Physical comorbidities and their relationship with cancer treatment and its outcomes in older adult populations: systematic review. JMIR cancer. 2021;7(4):e26425.

28. Pearce J, Martin S, Heritage S, et al. Frailty and outcomes in adults undergoing systemic anticancer treatment: a systematic review and meta-analysis. J Natl Cancer Inst. Jul 1 2025;117(7):1316–1339. doi:10.1093/jnci/djaf017

29. Mao Y, Xie H, Lv M, et al. The landscape of objective response rate of anti-PD-1/L1 monotherapy across 31 types of cancer: a system review and novel biomarker investigating. Cancer Immunol Immunother. Jul 2023;72(7):2483–2498. doi:10.1007/s00262-023-03441-3

30. Halle BR, Johnson DB. Defining and targeting BRAF mutations in solid tumors. Curr Treat Options Oncol. 2021;22(4):30.

31. Lovly CM, Salama AK, Salgia R. Tumor heterogeneity and therapeutic resistance. 2016:e585.

32. Chu AK, Kearns E, Holy C, et al. Cycling Through Uncertainty: The Psychological Experiences of Individuals With Lung Cancer Receiving Immunotherapy And/Or Targeted Therapy. Psychooncology. 2026/01/01 2026;35(1):e70372. 10.1002/pon.70372

33. Boulanger MC, Falade AS, Hsu K, et al. Patient and Caregiver Experience With the Hope and Prognostic Uncertainty of Immunotherapy: A Qualitative Study. JCO Oncol Pract. Feb 2025;21(2):178–187. doi:10.1200/op.24.00299

34. Gilligan T, Coyle N, Frankel RM, et al. Patient-Clinician Communication: American Society of Clinical Oncology Consensus Guideline. J Clin Oncol. Nov 1 2017;35(31):3618–3632. doi:10.1200/jco.2017.75.2311

35. Wong ML, Nicosia FM, Smith AK, et al. “You have to be sure that the patient has the full picture”: Adaptation of the Best Case/Worst Case communication tool for geriatric oncology. J Geriatr Oncol. Jun 2022;13(5):606–613. doi:10.1016/j.jgo.2022.01.014

36. Kiely B, McCaughan G, Christodoulou S, et al. Using scenarios to explain life expectancy in advanced cancer: attitudes of people with a cancer experience. Support Care Cancer. 2013;21(2):369–376.

37. Nahm SH, Stockler MR, Martin AJ, et al. Using three scenarios to explain life expectancy in advanced cancer: attitudes of patients, family members, and other healthcare professionals. Support Care Cancer. 2022;30(9):7763–7772.

38. Bauman JR, Temel JS. The integration of early palliative care with oncology care: the time has come for a new tradition. J Natl Compr Canc Netw. Dec 2014;12(12):1763–71; quiz 1771. doi:10.6004/jnccn.2014.0177

39. Greer JA, Jackson VA, Meier DE, Temel JS. Early integration of palliative care services with standard oncology care for patients with advanced cancer. CA Cancer J Clin. Sep 2013;63(5):349–63. doi:10.3322/caac.21192

40. Pilard C, Ancion M, Delvenne P, Jerusalem G, Hubert P, Herfs M. Cancer immunotherapy: it’s time to better predict patients’ response. Br J Cancer. Sep 2021;125(7):927–938. doi:10.1038/s41416-021-01413-x

41. Liu C, Yang M, Zhang D, Chen M, Zhu D. Clinical cancer immunotherapy: Current progress and prospects. Front Immunol. 2022;13:961805. doi:10.3389/fimmu.2022.961805

42. Zhai Z, Yu X, Yang B, et al. Colorectal cancer heterogeneity and targeted therapy: Clinical implications, challenges and solutions for treatment resistance. Semin Cell Dev Biol. Apr 2017;64:107–115. doi:10.1016/j.semcdb.2016.08.033

43. Rivera-Concepcion J, Uprety D, Adjei AA. Challenges in the Use of Targeted Therapies in Non-Small Cell Lung Cancer. Cancer Res Treat. Apr 2022;54(2):315–329. doi:10.4143/crt.2022.078

44. Parikh RB, Min EJ, Wileyto EP, et al. Uptake and Survival Outcomes Following Immune Checkpoint Inhibitor Therapy Among Trial-Ineligible Patients With Advanced Solid Cancers. JAMA Oncol. Dec 1 2021;7(12):1843–1850. doi:10.1001/jamaoncol.2021.4971

45. Krishnan M, Kasinath P, High R, Yu F, Teply BA. Impact of Performance Status on Response and Survival Among Patients Receiving Checkpoint Inhibitors for Advanced Solid Tumors. JCO Oncol Pract. Jan 2022;18(1):e175–e182. doi:10.1200/op.20.01055

46. Shah NJ, Della Pia A, Wu T, et al. Clinical Outcomes of Immune Checkpoint Inhibitors in Unique Cohorts Underrepresented in Clinical Trials. Cancers (Basel). Jun 14 2024;16(12)doi:10.3390/cancers16122223

47. Slotman E, de Munck L, Jager A, et al. Characteristics, treatment and survival in de novo and metachronous metastatic breast cancer: a nationwide comparative analysis. Breast Cancer Res Treat. 2026/05/12 2026;217(2):33. doi:10.1007/s10549-026-07979-5

