## Supplementary table 1 for "Real-world systemic therapy utilization and survival in synchronous metastatic solid cancer: a comprehensive nationwide analysis"

*Supplementary Table 1: Categorization of cancer types*

| **Cancer type** | **Description** | **Details** | **Topography codes** | **Morphology** |
| --- | --- | --- | --- | --- |
| HNSCC | Lip, Tongue, Gum, Floor of mouth, Palate, Other unspecified parts of mouth, Tonsil, Oropharynx, Nasopharynx, Pyriform Sinus, Hypopharynx, Other and ill-defines sites in lip, oral cavity and pharynx, larynx | Squamous cell carcinoma | C0.03-0.05, C01.0-01.9, C02.0-02.9, C03.0-03.9, C04.0-04.9, C05.0-05.9, C06.0-06.9, C09.0-09.9, C10.0-10.9, C11.0-11.9, C12.0-12.9, C13.0-13.9, C14.0-14.9, C32.0-32.9 | 8050-8089 |
| Esophageal/cardia cancer | Esophagus, Cardia NOS | Only carcinoma | C15.0-15.9, C16.0 | 8000-8576, 8940-8950, 8980-8983  Excluding NETs |
| Gastric cancer | Stomach | Only carcinoma | C16.1-C16.9 |  |
| Colon cancer | Colon | Only carcinoma | C18.0-18.9 |  |
| Rectal cancer | Rectosigmoid junction, Rectum | Only carcinoma | C19.9, C20.9 |  |
| Hepatocellular cancer | Hepatocellular carcinoma |  | C22.0 | 8000-8012, 8014-8040, 8046-8148, 8160-8231, 8250-8573, 8575-8576, 8980, 8982, 9110 |
| Gallbladder cancer | Gallbladder | Only carcinoma | C23.9 | 8000-8576, 8940-8950, 8980-8983 |
| Bile duct cancer | Intrahepatic bile duct, other and unspecified parts of biliary tract, biliary tract NOS | Only carcinoma | C22.1, C24.0-24.9 | 8000-8576, 8940-8950, 8980-8983  Excluding NETs |
| Pancreatic cancer | Pancreas | Only carcinoma | C25.0-C25.0 |  |
| NET | NET of the appendix |  | C18.1 | 8240-8242, 8248-8249 |
|  | NET of the stomach |  | C16.0-16.9 | 8240-8242, 8248-8249, 8150-8153, 8155-8156 |
|  | NET of the small intestine |  | C17.1-17.9 | 8240-8242, 8248-8249, 8150-8153, 8155-8156 |
|  | NET of the colon |  | C18.0, C18.2-18.9, C19.9, C20.9 | 8240-8242, 8248-8249 |
|  | Gastrointestinal NET other |  | C15.0-15.9, C26.0-26.9, C48.0-48.9 | 8240-8242, 8248-8249, 8150-8153, 8155-8156 |
|  | NET of the pancreas |  | C25.0-25.9 | 8240-8242, 8248-8249, 8150-8153, 8155-8157 |
|  | NET van duodenum/ampulla of Vater |  | C17.0, C24.1 | 8240-8242, 8248-8249, 8150-8153, 8155-8156 |
|  | NET with unknown primary location |  | C80.9 | 8240-8242, 8248-8249, 8150-8153, 8155-8156 |
| NSCLC | NSCLC |  | C34.0-34.9 | 8010-8020, 8022-8035, 8046-8230, 8243-8246, 8250-8576, 8972, 8980-8983 |
| SCLC | SCLC |  | C34.0-34.9 | 8041-8045, 8002, 8021 |
| Melanoma | Melanoma of the lip, skin, vulva, penis, scrotum NOS, unknown primary | Melanoma | C00.1-00.2, C44.0-44.9, C51,0-51.9, C60.0-60.9, C63.2, C80.9 | 8720-8790 |
| Sarcoma of the bone and soft tissues | Sarcoma of the soft tissues and viscera |  | C00-C39, C42, C44, C47-C52, C58-C69, C80 | 8710-8714, 8800-8830, 8833-8850, 8852-8921, 8935, 8963, 8990-8991, 9040-9045, 9120-9137, 9141-9221, 9230-9342, 9364-9373, 9540-9582 |
|  |  |  | C49 | 8000-8005, 8982 |
|  | Bones, joints, and articular cartilage |  | C40.0-40.9, C41.0-41.9 | 8000-8005, 8710-8714, 8800-8830, 8833-8850, 8852-8921, 8990-8991, 9040-9045, 9120-9137, 9141-9221, 9230-9342, 9364-9373, 9540-9582, 8935, 8963 |
| Breast cancer | Breast | Only carcinoma | C50.0-50.9 | 8000-8576, 8940-8950, 8980-8983, 9110 |
| Cervical cancer | Cervix Uteri carcinoma | Only carcinoma | C53.0-53.9 | 8000-8576, 8940-8951, 8980-8983, 9110 |
| Endometrial cancer | Corpus uteri, Uterus NOS | Endometrium carcinoma | C54.0-54.9, C55.9 |  |
| Ovarian cancer | Epithelial ovarian carcinoma,  Extra-ovarian carcinoma  Tubal carcinoma | Ovary | C56.0-56.9  C48.1-48.2  C57.0 | 8000-8239, 8250-8441, 8450, 8452-8461, 8470-8471, 8474, 8480-8576, 8930-8934, 8950-8951, 8980, 8982, 9000-9015, 9110  8000-8046, 8260-8576, 8950-8951, 8140, 8255, 8980, 9110  8000-8576, 8930-8934, 8950-8951, 9000-9015, 8980, 8982, 9110 |
| Prostate cancer | Prostate | Only carcinoma | C61.9 | 8000-8084, 8140-8576, 8940-8950, 8980-8983, 9110 |
| Kidney cancer | Kidney carcinoma | Only carcinoma | C64.0-64.9 | 8000-8084, 8140-8576, 8940-8950, 8980-8983, 9110 |
| Bladder cancer | Bladder, Renal pelvis, Ureter, Other and unspecified urinary organs | Only carcinoma | C67.0-67.9, C65.9, C66,9, C68.0-68.9  C64.0-64.9 | 8000-8576, 8940-8950, 8980-8983, 9110,  8120-8131 |
| Cancer of unknown primary | Unknown primary site |  | C80.9 | Excluding GIST/NET/melanomas specified above |
| Abbreviations: BCC: Basal cell carcinoma; GEP NET: Gastroenteropancreatic neuroendocrine tumor; GIST: Gastrointestinal stromal tumors; HNSCC: Head and neck squamous cell carcinoma; NET: neuroendocrine tumors; NSCLC: Non-small cell lung cancer; NOS: Not other specified; SCC: Squamous cell carcinoma; SCLC: Small cell lung cancer. | | | | |
