## Supplementary Table 2 for "Real-world systemic therapy utilization and survival in synchronous metastatic solid cancer: a comprehensive nationwide analysis"

*Supplementary Table 2: Categorization of systemic therapies*

**1. Immunotherapy**

| **NCR** | **LBZ add-on* medications** |
| --- | --- |
| Atezolizumab | Atezolizumab |
| Avelumab | Avelumab |
| Cemiplimab | Cemiplimab |
| Dostarlimab | Durvalumab |
| Durvalumab | Ipilimumab |
| Ipilimumab | Nivolumab |
| Ipilimumab/nivolumab | Pembrolizumab |
| Nivolumab |  |
| Nivolumab and relatlimab |  |
| Nivolumab/cabozantinib** |  |
| Paclitaxel/carboplatin/pembrolizumab |  |
| Paclitaxel/cisplatin/pembrolizumab |  |
| Pembrolizumab |  |
| Pembrolizumab/axitinib** |  |
| Pemetrexed/carboplatin/nivolumab/ipilimumab |  |
| Pemetrexed/carboplatin/pembrolizumab |  |
| Pemetrexed/cisplatin/pembrolizumab |  |
| Tremelimumab |  |
| Tremelimumab/durvalumab |  |
| Immunotherapy with study medication |  |
| Immunotherapy via dendritic cells |  |
| Immunotherapy via medication |  |
| Immunotherapy not otherwise specified |  |

**2. Targeted therapy**

| **NCR** | **LBZ add-on* medications** |
| --- | --- |
| Abemaciclib | Abemaciclib |
| Adagrasib | Afatinib |
| Afatinib | Alectinib |
| Alectinib | Alpelisib |
| Avapritinib | Amivantamab |
| Axitinib | Avapritinib |
| Bevacizumab | Axitinib |
| Binimetinib | Bevacizumab |
| Brigatinib | Binimetinib |
| Cabozantinib | Brigatinib |
| Capmatinib | Cabozantinib |
| Ceritinib | Ceritinib |
| Cetuximab | Cetuximab |
| Cobimetinib | Cobimetinib |
| Crizotinib | Crizotinib |
| Dabrafenib | Dabrafenib |
| Encorafenib | Encorafenib |
| Enfortumab vedotin | Enfortumab vedotin |
| Entrectinib | Entrectinib |
| Erlotinib | Erlotinib |
| Everolimus | Everolimus |
| Gefitinib | Gefitinib |
| Imatinib | Imatinib |
| Lapatinib | Lapatinib |
| Lenvatinib | Larotrectinib |
| Lorlatinib | Lenvatinib |
| Mobocertinib | Lorlatinib |
| Nintedanib | Nintedanib |
| Niraparib | Niraparib |
| Nivolumab/cabozantinib** | Olaparib |
| Olaparib | Olaratumab |
| Olaratumab | Osimertinib |
| Osimertinib | Palbociclib |
| Paclitaxel/carboplatin/bevacizumab | Panitumumab |
| Palbociclib | Pazopanib |
| Panitumumab | Pertuzumab |
| Pazopanib | Pertuzumab and trastuzumab |
| Pembrolizumab/axitinib** | Ramucirumab |
| Pertuzumab | Regorafenib |
| Pertuzumab and trastuzumab | Ribociclib |
| Poly(ADP-ribose) polymerase (PARP) inhibitor | Rucaparib |
| Pralsetinib | Selpercatinib |
| Protein kinase inhibitor | Sorafenib |
| Ramucirumab | Sunitinib |
| Ribociclib | Talazoparib |
| Selpercatinib | Tepotinib |
| Sirolimus | Trametinib |
| Sorafenib | Trastuzumab |
| Sotorasib | Trastuzumab deruxtecan |
| Targeted therapy with study medication | Trastuzumab emtansine |
| Sunitinib | Tucatinib |
| Temsirolimus | Vemurafenib |
| Tepotinib |  |
| Trametinib |  |
| Trastuzumab (Herceptin) |  |
| Trastuzumab deruxtecan |  |
| Trastuzumab emtansine |  |
| Veliparib |  |
| Vemurafenib |  |

**3. Novel hormonal agents & radionuclides**

**3.1 Novel hormonal agents**

| **NCR** | **LBZ add-on* medications** |
| --- | --- |
| Abiraterone | Abiraterone |
| Apalutamide | Apalutamide |
| Enzalutamide | Enzalutamide |

**3.2 Radionuclides**

| **NCR** | **LBZ add-on* medications** |
| --- | --- |
| Lutetium 177 lu oxodotreotide | Lutetium 177 lu oxodotreotide |
| PRRT (90Y-/177Lu-octreotate) | PRRT (90Y-/177Lu-octreotate) |
| Radium 223 ra dichloride | Radium 223 ra dichloride |

**4. Traditional systemic therapy**

**4.1 Chemotherapy**

| **NCR data** | **LBZ add-on* medications** | **LBZ treatment procedures** |
| --- | --- | --- |
| All chemotherapies | Aminolevulinic acid | Chemotherapy by IV or injection for metastatic or hematological tumors |
|  | Amsacrine | Chemotherapy not by IV or injection for metastatic or hematological tumors |
|  | Anagrelide | Providing guidance to an oncology patient during oral chemotherapy for metastases |
|  | Arsenic trioxid |  |
|  | Asparaginase |  |
|  | Azacitidine |  |
|  | Bendamustine |  |
|  | Bexarotene |  |
|  | Bleomycin |  |
|  | Busulfan |  |
|  | Cabazitaxel |  |
|  | Capecitabine |  |
|  | Chlorambucil |  |
|  | Chloromethine |  |
|  | Cisplatin |  |
|  | Clofarabine |  |
|  | Cyclophosphamide |  |
|  | Cytarabine |  |
|  | Cytarabine with Daunorubicin |  |
|  | Decitabine |  |
|  | Docetaxel |  |
|  | Doxorubicin |  |
|  | Epirubicin |  |
|  | Eribulin |  |
|  | Etoposide |  |
|  | Estramustine |  |
|  | Fludarabine |  |
|  | Gemcitabine |  |
|  | Hydroxycarbamide |  |
|  | Idarubicin |  |
|  | Irinotecan |  |
|  | Lomustine |  |
|  | Melphalan |  |
|  | Methylaminolevulinate |  |
|  | Mitomycin |  |
|  | Mitotane |  |
|  | Mitoxantrone |  |
|  | Nelarabine |  |
|  | Oxaliplatin |  |
|  | Paclitaxel |  |
|  | Pegaspargase |  |
|  | Panobinostat |  |
|  | Pemetrexed |  |
|  | Pixantrone |  |
|  | Procarbazine |  |
|  | Temoporfin |  |
|  | Temozolomide |  |
|  | Teniposide |  |
|  | Tegafur, combination preparations |  |
|  | Thiotepa |  |
|  | Topotecan |  |
|  | Trabectedin |  |
|  | Trifluridine, combination preparations |  |
|  | Tretinoin |  |
|  | Vinblastine |  |
|  | Vincristine |  |
|  | Vinorelbine |  |

**4.2 Traditional hormone therapy**

| **NCR data** | **LBZ treatment procedures***** |
| --- | --- |
| All hormonal therapies except the add-on hormone therapies | Hormonal therapy by IV or injection for metastatic or hematological tumors |
|  | Hormonal therapy not by IV or injection for metastatic or hematological tumors |
|  | Providing guidance to an oncology patient during hormonal therapy for metastases |
|  | Injection of LHRH (Luteinizing Hormone-Releasing Hormone) analogue/antagonist |

* Add-on medications are high-cost medicines that are reimbursed separately from other hospital care in the Netherlands

** Excluded in the sensitivity analyses, where immunotherapy and targeted therapy were analyzed as mutually exclusive groups

*** Patients with these treatment procedures who have also had a novel hormonal agent (an add-on hormonal medication) were excluded
