## Supplementary Table 3 for "Real-world systemic therapy utilization and survival in synchronous metastatic solid cancer: a comprehensive nationwide analysis"

*Supplementary Table 3: Characteristics of all patients diagnosed with the novo metastatic cancer in 2018-2022 and those who were linked to the Dutch National Hospital Care Registration (LBZ)*

|  | **Total cohort 2018-2022** | | | **Patients linked to the LBZ** | | |
| --- | --- | --- | --- | --- | --- | --- |
| **Cancer type** | **N** | **Median age (IQR)** | **% female** | **N** | **Median age (IQR)** | **% female** |
| Total cohort | 102947 | 71 (63-78) | 42 | 82544 | 71 (62-77) | 42 |
| Bile duct cancer | 1906 | 70 (63-77) | 50 | 1556 | 70 (62-77) | 50 |
| Bladder cancer | 2841 | 73 (67-80) | 33 | 2235 | 73 (66-80) | 33 |
| Breast cancer | 4882 | 65 (52-75) | 99 | 3942 | 64 (52-75) | 99 |
| Cervical cancer | 338 | 56 (48-70) | 100 | 298 | 56 (48-70) | 100 |
| Colon cancer | 10331 | 72 (61-79) | 49 | 8264 | 71 (61-79) | 48 |
| Endometrial cancer | 1033 | 72 (64-77) | 100 | 857 | 71 (63-77) | 100 |
| Esophageal/cardia cancer | 5459 | 70 (62-76) | 23 | 4513 | 70 (62-76) | 23 |
| Gallbladder cancer | 570 | 72 (64-79) | 66 | 474 | 72 (63-78) | 65 |
| Gastric cancer | 3236 | 71 (61-78) | 39 | 2635 | 71 (61-78) | 39 |
| HNSCC | 416 | 69 (61-75) | 25 | 383 | 69 (61-75) | 25 |
| Hepatocellular cancer | 721 | 72 (65-77) | 23 | 597 | 72 (65-77) | 23 |
| Kidney cancer | 2607 | 70 (62-77) | 32 | 2231 | 70 (62-76) | 31 |
| Melanoma | 1525 | 68 (57-76) | 36 | 1369 | 68 (57-76) | 34 |
| NET | 1628 | 68 (59-75) | 44 | 1405 | 68 (59-75) | 44 |
| NSCLC | 25512 | 69 (62-75) | 45 | 20772 | 69 (62-75) | 45 |
| Ovarian cancer | 2209 | 71 (62-78) | 100 | 1600 | 71 (61-77) | 100 |
| Pancreatic cancer | 8434 | 72 (64-78) | 49 | 6499 | 72 (64-78) | 49 |
| Prostate cancer | 13684 | 74 (68-80) | 0 | 10726 | 74 (68-80) | 0 |
| Rectal cancer | 3402 | 68 (57-77) | 38 | 2824 | 68 (57-77) | 38 |
| SCLC | 5683 | 69 (63-75) | 50 | 4566 | 69 (63-75) | 50 |
| Sarcoma | 750 | 66 (52-75) | 40 | 542 | 64 (52-74) | 40 |
| Cancer of unknown primary | 5780 | 76 (68-83) | 49 | 4256 | 76 (68-83) | 49 |
