## Supplementary Table 4 for "Real-world systemic therapy utilization and survival in synchronous metastatic solid cancer: a comprehensive nationwide analysis"

*Supplementary Table 4: Median follow-up (FU) in months of patients diagnosed in 2018-2022 and linked to the LBZ, stratified by type of systemic treatment*

|  | **Immunotherapy** | | **Targeted therapy** | | **Novel hormonal agents and/or radionuclides** | | **Traditional systemic therapy only (chemotherapy, hormone therapy)** | |
| --- | --- | --- | --- | --- | --- | --- | --- | --- |
| **Cancer type** | **N** | **Median FU (months)** | **N** | **Median FU (months)** | **N** | **Median FU (months)** | **N** | **Median FU (months)** |
| Total | 12069 | 63 | 12117 | 67 | 4125 | 69 | 23903 | 63 |
| Bile duct cancer | 8 | x | 10 | x | 0 | x | 556 | 57 |
| Bladder cancer | 405 | 61 | 31 | x | 3 | x | 571 | 69 |
| Breast cancer | 74 | 53 | 1757 | 69 | 0 | x | 1750 | 61 |
| Cervical cancer | 19 | x | 83 | 55 | 0 | x | 82 | 52 |
| Colon cancer | 147 | 53 | 3064 | 67 | 6 | x | 1567 | 66 |
| Endometrial cancer | 21 | x | 23 | x | 0 | x | 552 | 64 |
| Esophageal/cardia cancer | 143 | 43 | 684 | 62 | 1 | x | 1580 | 64 |
| Gallbladder cancer | 1 | x | 1 | x | 1 | x | 176 | 46 |
| Gastric cancer | 54 | 44 | 369 | 78 | 0 | x | 844 | 67 |
| HNSCC | 69 | 65 | 34 | x | 2 | x | 32 | x |
| Hepatocellular cancer | 59 | 58 | 160 | 58 | 0 | x | 6 | x |
| Kidney cancer | 991 | 64 | 846 | 68 | 1 | x | 12 | x |
| Melanoma | 880 | 61 | 389 | 68 | 6 | x | 8 | x |
| NET | 11 | x | 89 | 80 | 189 | 73 | 716 | 64 |
| NSCLC | 8855 | 64 | 2524 | 65 | 8 | x | 1961 | 78 |
| Ovarian cancer | 37 | x | 474 | 64 | 0 | x | 764 | 69 |
| Pancreatic cancer | 29 | x | 29 | x | 2 | x | 2024 | 59 |
| Prostate cancer | 47 | x | 171 | 74 | 3899 | 68 | 6316 | 62 |
| Rectal cancer | 15 | x | 1214 | 71 | 3 | x | 686 | 62 |
| SCLC | 116 | 80 | 30 | x | 0 | x | 3105 | 70 |
| Sarcoma | 7 | x | 69 | 61 | 1 | x | 156 | 57 |
| Cancer of unknown primary | 81 | 72 | 66 | 70 | 3 | x | 439 | 65 |

x –Treatment groups with fewer than 50 patients were excluded from presentation
