## Supplementary Figures 1 and 2 for "Real-world systemic therapy utilization and survival in synchronous metastatic solid cancer: a comprehensive nationwide analysis"

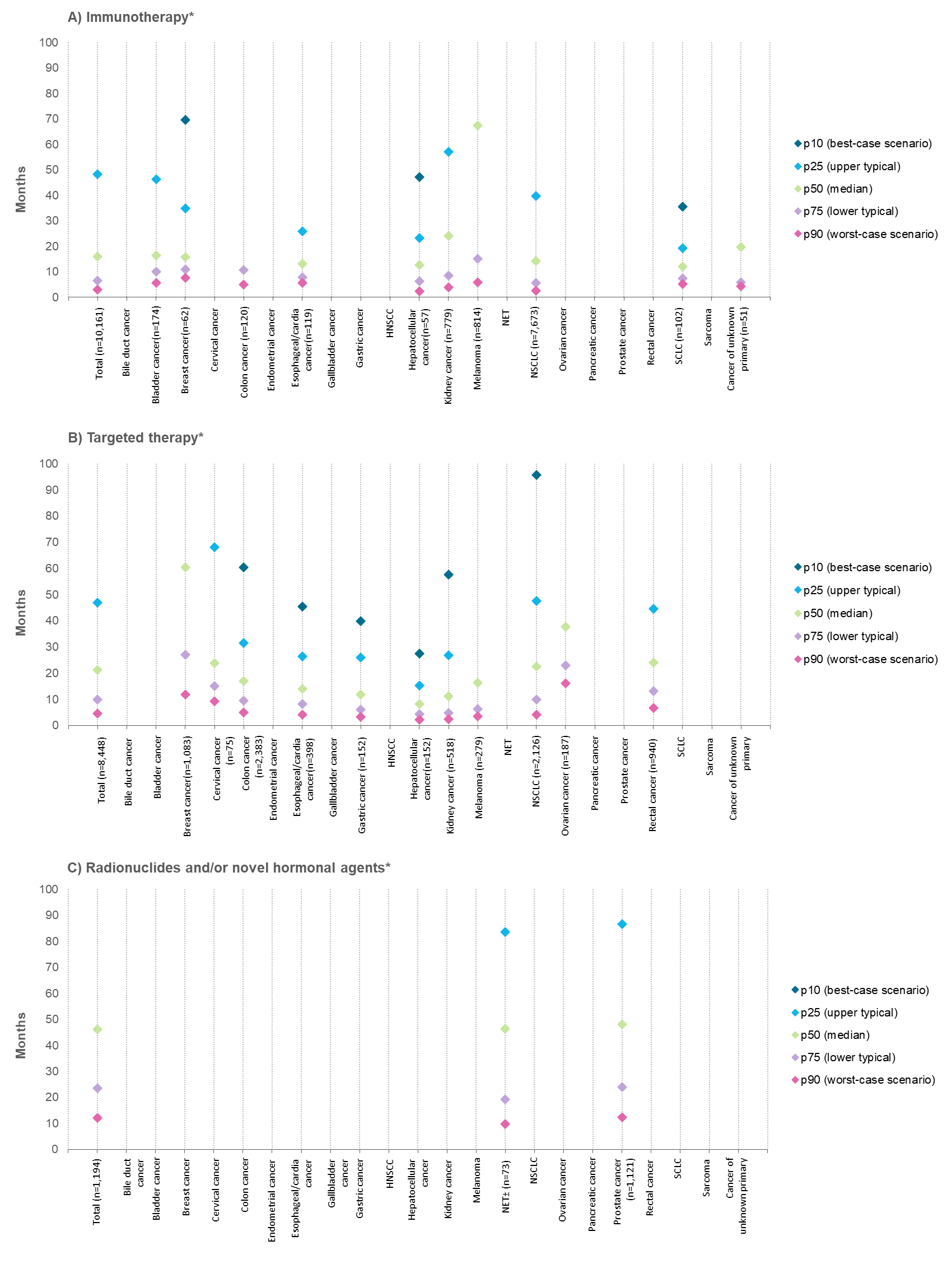

*Supplementary Figure 1: Survival scenarios in patients diagnosed with synchronous metastatic cancer in* ***2018-2022*** *treated with novel systemic anticancer therapy as initial treatment.*

**Categories are not mutually exclusive.*

*± In NET only radionuclides were administered.*

*Notes:*

*- Systemic treatment data were available from January 1, 2018, until December 31, 2023.*

*- For some treatment groups and cancer types, the highest percentiles cannot be reported due to insufficient follow-up time to calculate point estimates.*

*- Treatment groups with fewer than 50 patients were excluded from presentation*

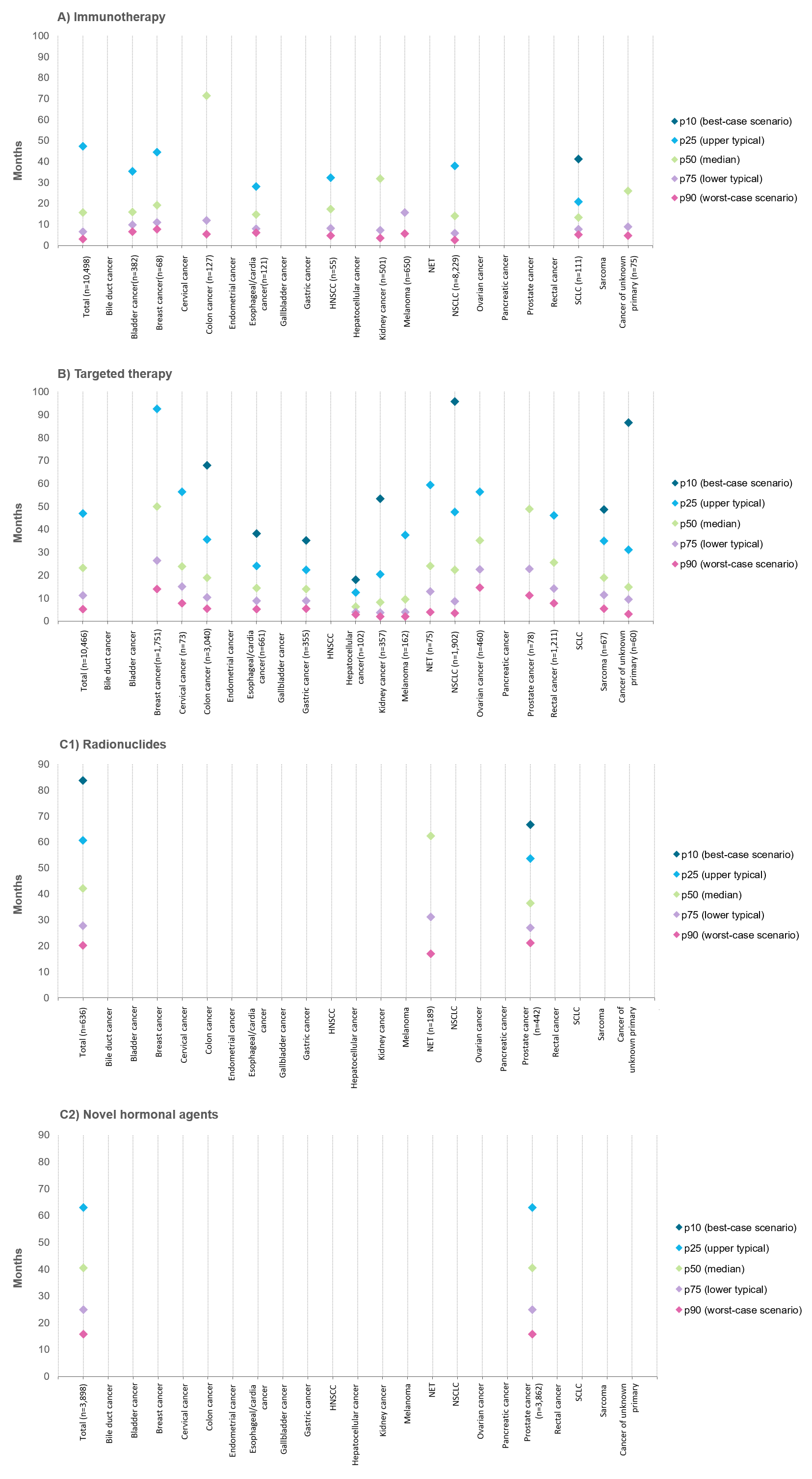

*Supplementary Figure 2: Survival scenarios in patients diagnosed with synchronous metastatic cancer in* ***2018-2022*** *treated with only one type of novel systemic anticancer therapy (with or without traditional systemic therapy).*

*Notes:*

*- Systemic treatment data were available from January 1, 2018, until December 31, 2023.*

*- For some treatment groups and cancer types, the highest percentiles cannot be reported due to insufficient follow-up time to calculate point estimates.*

*- Treatment groups with fewer than 50 patients were excluded from presentation*
